# Learning to fake it: limited responses and fabricated references provided by ChatGPT for medical questions

**DOI:** 10.1101/2023.03.16.23286914

**Authors:** Jocelyn Gravel, Madeleine D’Amours-Gravel, Esli Osmanlliu

## Abstract

**Background:** ChatGPT have gained public notoriety and recently supported manuscript preparation. Our objective was to evaluate the quality of the answers and the references provided by ChatGPT for medical questions.

**Methods:** Three researchers asked ChatGPT a total of 20 medical questions and prompted it to provide the corresponding references. The responses were evaluated for quality of content by medical experts using a verbal numeric scale going from 0 to 100%. These experts were the corresponding author of the 20 articles from where the medical questions were derived. We planned to evaluate three references per response for their pertinence, but this was amended based on preliminary results showing that most references provided by ChatGPT were fabricated.

**Results:** ChatGPT provided responses varying between 53 and 244 words long and reported two to seven references per answer. Seventeen of the 20 invited raters provided feedback. The raters reported limited quality of the responses with a median score of 60% (1^st^ and 3^rd^ quartile: 50% and 85%). Additionally, they identified major (n=5) and minor (n=7) factual errors among the 17 evaluated responses. Of the 59 references evaluated, 41 (69%) were fabricated, though they appeared real. Most fabricated citations used names of authors with previous relevant publications, a title that seemed pertinent and a credible journal format.

**Interpretation:** When asked multiple medical questions, ChatGPT provided answers of limited quality for scientific publication. More importantly, ChatGPT provided deceptively real references. Users of ChatGPT should pay particular attention to the references provided before integration into medical manuscripts.

## Introduction

Large Language Models (LLM) constitute a branch of artificial intelligence (AI) at the intersection of linguistic and computer science(1). Trained on massive quantities of text-based data, LLMs have learned to interpret written input and produce language that is understandable to humans. LLMs incorporate several algorithms, including generative pre-trained transformers (GPT). This type of neural network architecture is useful in chatbots, rendering them particularly effective at simulating human conversations. On November 2022, San Francisco-based company OpenAI released a freely available version of ChatGPT, a LLM based on their proprietary GPT (GPT-3)(2). Since then, scientific articles have been written in part by ChatGPT and published(3-5). These publications were mainly to demonstrate the remarkable quality of the manuscripts written by ChatGPT. For example, a journal published an editorial for which the first five paragraphs were written by ChatGPT(4). In another publication, the researcher asked ChatGPT to discuss the potential impact of taking Rapamycin to increase longevity(5). The chatbot’s output constituted most of the article. Finally, an article describing the potential use of ChatGPT for medical writing was mainly written by the chatbot(3). In these articles, the parts produced by the chatbot did not include any references. While being low, the number of publications discussing ChatGPT has exploded in the last weeks. It went from only 24 publications identified in Pubmed with the term “ChatGPT” on February 7^th^, 2023, to 92 publications on March 6^th^. These publications were mainly editorials or news from scientific journals praising the quality of the writing (1, 6-8) or questioning ethical aspects of using chatbot for scientific writing(9-12).

While most articles suggest that the answers provided by ChatGPT are acceptable, there is no information about the sources of its knowledge and no reference is usually provided by ChatGPT, unless specifically requested. This limitation is acknowledged by OpenAI as the model was trained on multiple internet texts with a large number of sources(2). However, when conversing with ChatGPT, one can prompt it regarding the references backing its assertions.

To our knowledge, no study has evaluated the quality and appropriateness of the references provided by ChatGPT. This study aimed to evaluate the quality of the answers and corresponding references provided by ChatGPT, when responding to a wide range of medical questions. A secondary objective arose during the construction of the study as many citations provided by ChatGPT were not found. Because of this, we aimed to assess the validity of the suggested references.

## Methods

This was an experimental observational study conducted in February 2023 evaluating the quality of the responses provided by ChatGPT (Version 3.5. OpenAI Inc, San Francisco, CA, USA) to 20 medical questions from diverse fields.

The medical questions were identified by selecting five research articles published at the end of 2022 in four high-impact factor medical journals (BMJ(13-17), CMAJ(18-22), the Lancet(23-32) and NEJM). These 20 articles spanned different topics and fields. The questions asked to ChatGPT were related to the primary objectives of the 20 studies. In most cases, it was the stated primary objective of the study. In a few instances where the objective was deemed too narrow to ensure a minimal breadth of references, we formulated the question in a broader context. For example, the primary objective “*to critically examine the leadership experiences of African Nova Scotian nurses in health care systems*?”(18), was changed to: *What are the leadership experiences of African nurses in the United States health care systems*?

In general, questions to ChatGPT started as “*What are…” (see example above)*, without any word limit or other constraint. Following the answer by ChatGPT, a follow-up question asked: *Do you have references for this*? All references were counted, but only the first three were used for the analysis. To promote the external validity of the study and considering that ChatGPT is sensitive to previous chats, questions were asked by the three co-authors on different computers and ChatGPT accounts.

The primary outcomes were: 1. The appropriateness of the references; 2. The quality of the responses. We initially aimed to evaluate the appropriateness of the references by multiple raters using the following verbal numeric scale: *On a scale of 0 to 100% where 0 signifies that the references had no relationship with the study question and 100% is for the three most pertinent references for this topic, how would you rate the references provided*? Our initial plan was to provide the articles to the raters. However, given that we failed to find the first six articles, we modified this outcome to evaluate whether the reference existed. To verify this, we searched Pubmed using the title and authors. If unsuccessful, we then searched in the journal’s website. To better describe the references provided by ChatGPT, we evaluated whether the authors listed had previous publication in the field. We also looked at the title (*Is it a title that seems pertinent for the study question?)* and the journal (*Does the citation look a plausible article for this journal?)*. The quality of the response was measured using the following verbal numeric scale from 0 to 100%: *On a scale of 0 to 100% where 0 is equal to no answer, 100% to a perfect answer and 50% to the minimum acceptable answer how would you rate the answer provided to the question*? Also, raters were asked to report any factual error found in the ChatGPT answer, as well as any other relevant qualitative feedback.

To ensure domain expertise for the selected articles, we invited the corresponding author of each article to act as rater and determine the quality of the response. We contacted each corresponding authors by email, inviting them to provide feedback on the answer related to their respective study objective. When the corresponding author did not reply, we contacted other listed authors. Each corresponding author was contacted at least three times before saying it was unsuccessful. These authors were deemed content experts in their field of publication.

The primary analysis of this study determined the validity of the references by calculating the proportion of references that really existed among all evaluated references. We also reported the proportion of references that listed an author with previous publications in the field of interest. The other primary analysis pertained to the quality of ChatGPT responses, reported as the median and interquartile range on the verbal numeric scale. We also reported the proportion of responses that contained factual errors according to the raters. Minor errors referred to erroneous details with limited impact to the quality of the overall response (e.g. an overoptimistic affirmation that is not supported by any data), whereas major errors consisted of flagrant mistakes that invalidated the response (e.g. wrong pathophysiological explanation).

We did not conduct the analysis using the verbal numeric scale for the appropriateness of the references due to the small number of real references.

We had no prespecified idea of the median scores that would be obtained for the responses. It was estimated that the evaluation of at least 12 questions would provide a great variety of study subjects and allows to demonstrate the general quality of the responses/references. Also, this would lead to at least 30 references to evaluate. Based on this, and the premise that at least 60% of the invited authors would agree to help us, we invited 20 authors to rate 20 study questions in order to have at least 12 evaluations.

We did not seek institutional review board approval given that all data were publicly available, and no participants were involved. The study was completed without financial support. The manuscript is an honest, accurate, and transparent account of the study being reported; no important aspects of the study have been omitted. Any discrepancies from the study as originally planned was explained (change in the outcome regarding the validity of the references).

## Results

Each of the 20 study questions were asked to ChatGPT by a member of the study team (JG n= 10, MDG n=5 and EO n=5) and received a response that varied in length between 53 and 309 words. When prompted to provide its references, ChatGPT provided two to seven references. A total of 59 references were included in the primary analysis.

When searching for the references suggested by ChatGPT, we noted that while they looked credible (see figure 1), most of them were fabricated by ChatGPT. Indeed, 56/59 (95%) references contained authors with previous publications on a related topic found in Pubmed or were from recognized organizations (ex: CDC, FDA, etc.) (see table 1). Also, all titles seemed appropriate because it was related to the study question. In reality, 41/59 (69%) references were fabricated. Among the 18 real references, 11 were titles of real published articles (including three with minor citation errors and five with major citation errors), five were existing websites and two were books (Figure 2). The remaining reference (n=41) did not exist. Of those, 29 (71%) of the fabricated articles were reportedly published in a known medical journal, website or manuscript repository (ex: CDC or MedRxiv) using an appropriate format of citation (i.e.: they reported a year, volume number and page that were coherent with the journal). However, the reported volume and page range pertained to an unrelated article. All responses and references can be found on the web appendix.

**Table 1.**
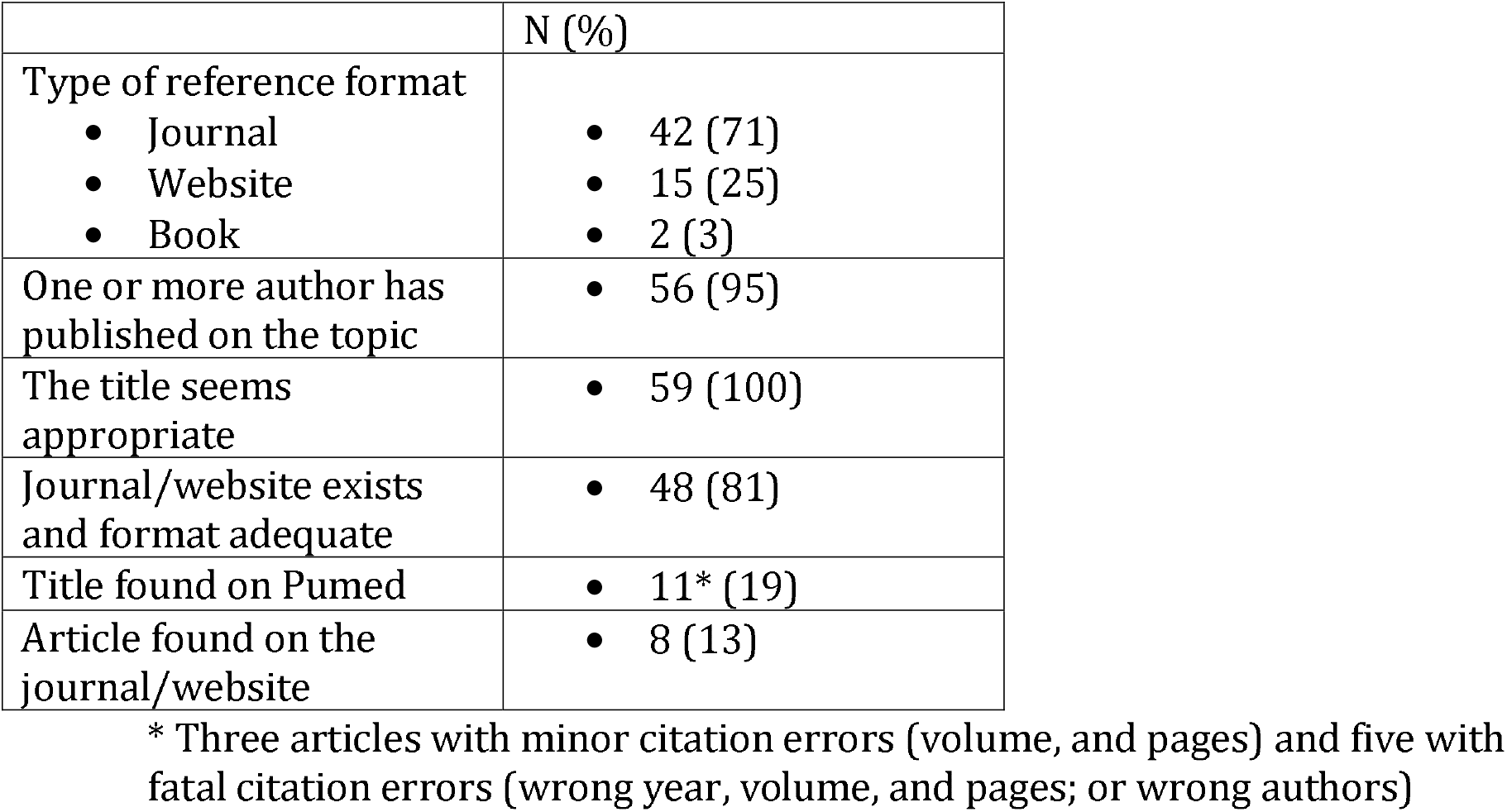
Characteristics of the references provided by ChatGPT (n=59)

**Figure 1.**
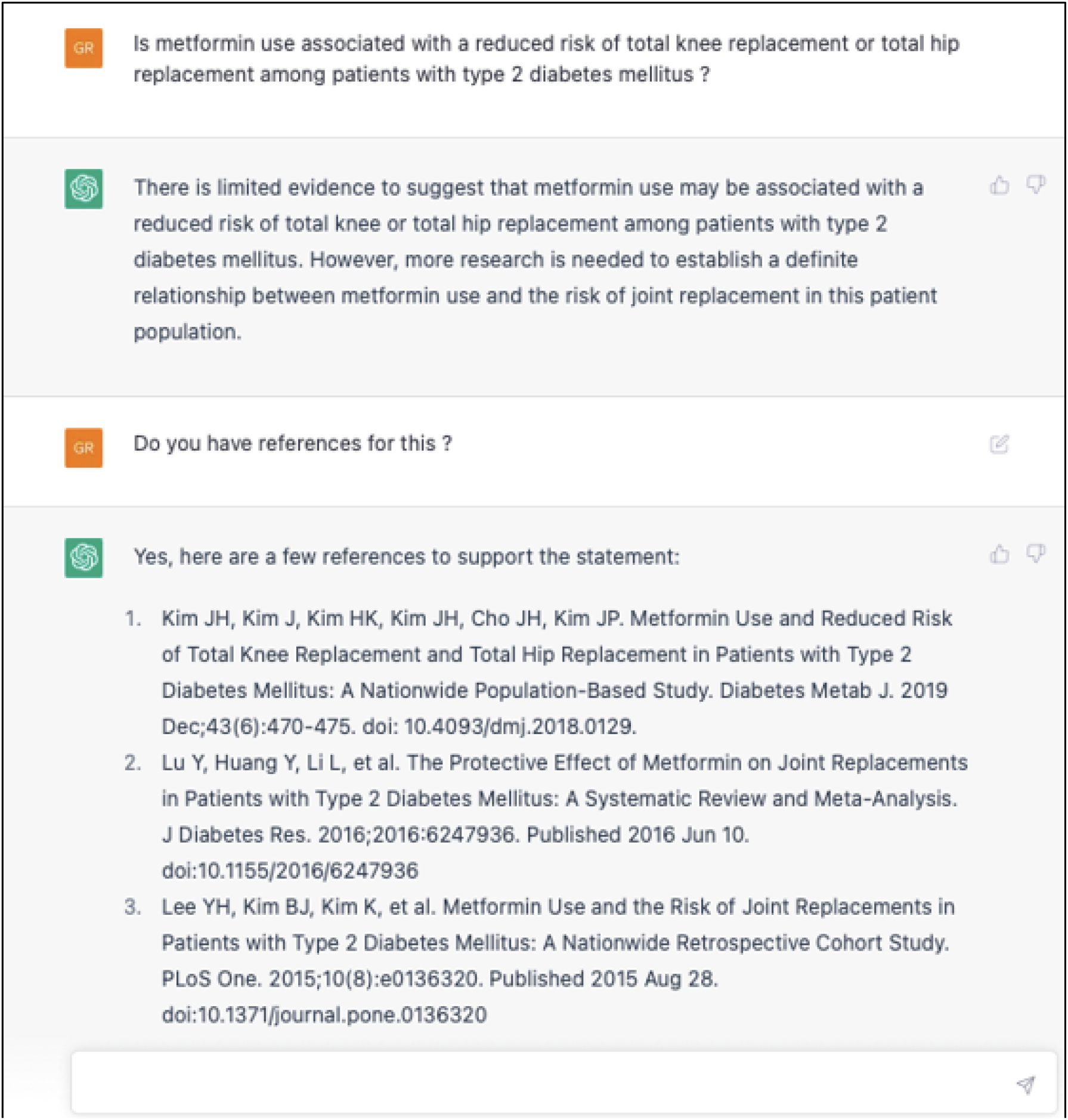
screenshot of an example of responses provided by ChatGPT

**Figure 2.**
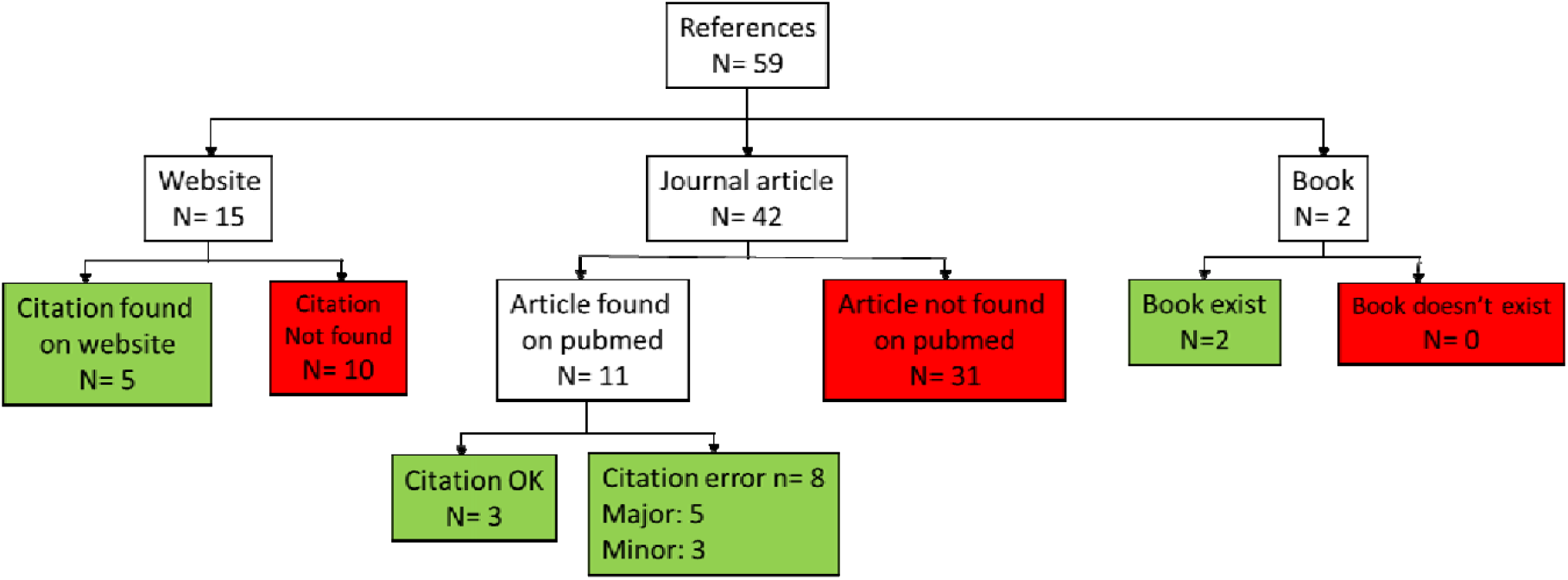
Distribution of the references provided by ChatGPT (n=59)

Of the 20 corresponding authors, 17 (0.85) agreed to evaluate the responses. The median score they provided was 60% (1^st^ and 3^rd^ quartiles 50% and 85%). Raters identified major factual error in five (0.29) responses. For example, a rater identified that the mechanism of action of antipsychotic described by ChatGPT was incorrect. Another noted that ChatGPT overestimated the global burden of mortality associated with Shigella infections by a factor of 10. Minor errors were identified in seven (0.41) other responses.

## Interpretation

This study demonstrated that over two-thirds of the references provided by ChatGPT to a diverse set of medical questions were fabricated, though most seemed deceptively real. Moreover, domain experts identified major factual errors in a quarter of the responses. These findings are alarming, given that trustworthiness is a pillar of scientific communication,

A previous study evaluated answers provided by ChatGPT using a scientific methodology in January 2023(33). This study demonstrated that ChatGPT’s score (60%) was lower than that of Korean medical students (90%) in a single parasitology examination. Another study compared results of ChatGPT to two other LLMs on the United States Medical Licensing Examination Step 1 and Step 2 exams using two question banks. With a mean score of 60%, they concluded that ChatGPT outperformed the other chatbots and “achieved the equivalent of a passing score for a third-year medical student”(34). A recent comment published in Nature reported that “ChatGPT fabricated a convincing response that contained several factual errors”(35). To our knowledge, this is the first study evaluating the quality of the references provided by ChatGPT. However, a recent article described a manuscript written by ChatGPT using mock data(36). When asked to conduct a literature search, ChatGPT suggested 9 references. The author of the article acknowledged that: “*Interestingly, at least some of these references that ChatGPT suggested do not exist in the form that is presented here*”. We looked for these 9 articles and, none exist. Reference inaccuracies do not constitute a novelty in medical publishing, but usually relate to minor mistakes(37-40). For example, Browne noted that most of the errors in referencing among papers submitted to any of five radiology journals were related to a failure to follow author submission guideliness(37). Another study reported that 15% of articles published in a single journal had an error but it was most commonly a spelling or punctuation error(38). Finally, two studies reported that approximately 15% of articles had errors, and up to 4.5% consisted of major errors. However, these errors did not relate to purely fabricated references.

The importance of proper referencing is undeniable. As suggested by Glick, “you are what you cite”(41). The quality and breadth of the references provided demonstrates that the researchers have done a complete literature review and are knowledgeable about the topic. This process enables the integration of findings in the context of previous work, a fundamental aspect of medical research advancement(42). It limits the risk of biases.

Failing to provide references is one thing but creating fake references would be considered fraudulent for researchers. When asked for references, ChatGPT made very appealing suggestions. It blended authors with a good research track to an interesting title in addition to a relevant journal like if the chatbot wanted to put the best of everything in a single reference. Some titles seemed to be the perfect article for our question. For example, for the question “*What is the impact of haloperidol in intensive care unit patients with delirium?”*, ChatGPT suggested the following inexistent title as reference: *“Haloperidol in critically ill patients with delirium: a randomized, placebo-controlled trial”*. In most cases, it suggested authors that published multiple scientific articles on the subject of interest.

This study highlights an important shortcoming of LLMs, namely their risk of being “confidently wrong”. While such models have now reached astounding performance in simulating human conversations, the next stages in their development must improve information validity and responsible deployment. Though a series of disclaimers upon user registration highlight some of the limitations of ChatGPT(2), scientists considering the use of this tool to support manuscript preparation must be aware of its limitations. To promote responsible responses, chatbot developers should adapt the reward models that guide the algorithm’s output, so that they learn to optimize the validity of the information and references provided. OpenAI uses a supervised learning framework described as reinforcement learning from human feedback(2), which should be amenable to such modifications.

These findings can help orient the use of LLMs by scientists in ways that are safe, contribute to their wellbeing and that of their teams, while targeting feasible applications(43). This includes the partial automation of tedious tasks, such as the initial steps in knowledge mapping and synthesis. This would free up time to critically appraise the information presented, thoroughly verify the corresponding sources, and orient subsequent searches. Future LLM iterations may also support manuscript formatting according to journal requirements, and facilitate knowledge translation to diverse audiences (e.g., translation, summarization and adjustment of readability). On the long term, use of LLM might improve knowledge translation by accelerating manuscript redaction and improve the quality of the writing. However, it could also threaten the scientific validity of manuscript if it incorporates inaccurate information and is misused(43). Researchers using ChatGPT may be misled by false information because clear, seemingly coherent and stylistically appealing references can conceal poor content quality. Journals should consider clear guidelines regarding the allowed uses and reporting guidelines when tools such as ChatGPT are used. In the future, such tools may be re-designed to support human reviewers and publishers when appraising submitted articles and their corresponding bibliography. This use case highlights the enormous potential and alarming pitfalls of LLM integration in scientific writing. The inherent risk of overconfidence among LLMs also emphasizes the need for “humans in the loop” as a key element for their responsible implementation.

There are limitations to this study. First, we always asked the same question regarding references and did not specify to ChatGPT to limit references to published articles. However, when trying this *a posteriori*, we obtained similar answers. Second, the information provided was accurate as of February 2023 and may improve in the following months/years. For example, there were two questions related to the COVID 19 pandemic. ChatGPT being constructed based on data published before sept 2021, it may have been difficult to find sufficient knowledge for adequate answers. Also, ChatGPT has been launched publicly in November 2022, with the stated objective of iterative cycles of learning and improvement. It is probable that LLM will improve and learn to provide responses exempt from factual error. Finally, we used multiple raters to measure the quality of the responses. We preferred to have experts in each specific field than to have experts in scoring.

## Conclusion

ChatGPT proposes undeniable progress. This study should alert the scientific community to be careful about the important risks of relying on its references because it is assembling very convincing, yet often fabricated citations. To be useful for medical editing, chatbots should embrace the values of the scientific community such as integrity and completeness. Considering the speed of improvement of LLMs, we are hopeful that future versions of ChatGPT will suggest more accurate responses when asked to provide references.

## Data Availability

All data produced in the present study are available upon reasonable request to the authors

## Acknowledgements

The study team would like to acknowledge the contribution of Drs Changhai Ding, Areef Ishani, Yazdan Yazdanpanah, Nina Christine Andersen-Ranberg, Marc Rothenberg, Evan S Dellon, Ken Parhar, Jason Weatherald, Alberto Ruano Raviña, John Cleland, John H Krystal, Luciana Vercoza Viana, Kevin Ituka, Miriam Sander, Amanda Roberts, Fan Wang and all many other authors who agreed to rate the quality of the responses for this study.

## Notes

**Financial Disclosure Statement:** The authors have no financial relationships relevant to this article to disclose. This study was conducted without financial support.

**Conflict of Interest:** The authors have no conflict of interest relevant to this article to disclose.

### Competing Interest Statement

The authors have declared no competing interest.

### Funding Statement

This study did not receive any funding

